# Identifying characteristics and clinical conditions associated with hand grip strength in adults: the Baseline Health Study

**DOI:** 10.1101/2023.02.16.23286051

**Authors:** Kenneth A. Taylor, Megan K. Carroll, Sarah Short, Adam P. Goode

## Abstract

**Background:** Low hand grip strength (HGS) predicts several conditions, but its value outside of the older adult population is unclear. We sought to identify the most salient factors associated with HGS using a rich list of candidate variables while stratifying by age and sex.

**Methods:** We used data from the initial visit from the Project Baseline Health Study (N=2502) which captured detailed demographic, occupational, social, lifestyle, and clinical data. We applied MI-LASSO using group methods to determine variables most associated with HGS out of 175 candidate variables. We performed analyses separately for sex and age (<65 vs. ≥65).

**Results:** Race was associated with HGS to varying degrees across groups. Osteoporosis and osteopenia were negatively associated with HGS in female study participants. Immune cell counts were negatively associated with HGS for male participants ≥65 (neutrophils) and female participants (≥65, monocytes; <65, lymphocytes). Most findings were age and/or sex group-specific; few were common across all groups.

**Conclusions:** Several of the variables associated with HGS in each group were novel, while others corroborate previous research. Our results support HGS as a useful indicator of a variety of clinical characteristics; however, its utility varies by age and sex.

## Introduction

Hand grip strength (HGS) is a noninvasive and reliably measured biomarker that is most commonly assessed with a hydraulic hand dynamometer. Primarily studied in older adults, HGS affords a consistent indicator of overall strength. However, it is most informative when paired with a measure of lower extremity strength [1]. Lower HGS in older adults is associated with limitations in functional mobility and upper extremity activity [2-6]. Likewise, decreased HGS in this population is also a consistent indicator of disease status and mental health status. Lower HGS is associated with conditions such as prediabetes[7], diabetes (especially with neuropathy), and their clinical indicators (e.g., higher hemoglobin A1c; higher fasting blood glucose) [8-11]. It is also associated with multimorbidity [12-15], malnutrition [16-19], sleep impairment [20-23], and psychological conditions such as depression [24-29] and impaired cognition [30,31].

Among healthy adults, HGS is highest in their 20s, typically maintaining through their 40s and decreasing thereafter, with consistent differences between sexes [32-41]. Using age-appropriate reference values, some researchers have identified cut points intended to identify older individuals with sarcopenia or clinically relevant weakness, and to predict disease or mortality [42-47]. For example, older adults with HGS values below 16 kg for females or 26-27 kg for males, may meet diagnostic criteria for sarcopenia and/or frailty and would benefit from further clinical assessment [35,42,48]. These cut points are generalizable to Japan, North America, Europe, and Australia, but not to other regions [49].

HGS also predicts future injury, disease, and mortality in older adults [50]. It is a strong predictor of incident cardiovascular disease, with estimated risk increasing 21% for each 5-kg decrease in HGS [51], although it may not predict specific incident cardiovascular events [52]. However, an increased risk of cardiovascular mortality is associated with lower HGS [51,53,54], with one study finding HGS to be a stronger predictor than systolic blood pressure for cardiovascular mortality, and other disease-specific and all-cause mortality [55]. This additional prognostic value of lower HGS as a predictor of all-cause mortality has been reported across several cohort studies from various geographical regions [51,53,56-66]. Low HGS also predicts future function in care-seeking and non-care-seeking populations [67-74] as well as falls and osteoporotic fractures among older adults [75-82], with the latter likely being directly related to reported associations with physical strength, functional mobility, and bone mineral density.

Current evidence suggests HGS as a biomarker with potential value for screening, diagnosis, and determining prognosis for a wide array of conditions. However, its value as an indicator/predictor of outcomes has not been well studied outside of the older adult population. Studies have typically focused on investigating HGS alongside other variables as indicators/predictors of a specific outcome rather than comprehensively investigating clinical factors and conditions associated with HGS.

The Project Baseline Health Study (PBHS) is a prospective cohort study of U.S. adults that has collected a wide array of personal, behavioral, and clinical characteristics that together comprise data sufficiently rich to allow deep phenotyping of study participants [83,84]. In this analysis, we examined participants in the BHS study cohort to identify the most salient factors associated with HGS by examining demographic, socioeconomic status-related health behaviors, medical conditions, symptoms, patient-reported outcomes, and laboratory measures across a wide adult age group while stratifying by age and sex to account for expected differences between these groups.

## Results

Of the 2502 participants in the deeply phenotyped BHS cohort, 2366 (95%) completed baseline HGS assessment; 1318 (55.7%) were female and 1048 (44.3%) were male. In this cohort, 63.5% of participants identified as White (64.2% female; 62.6% male), 16.1% as Black (16.7%; 15.3%), 10.3% as Asian (8.3%; 12.8%), and 8.0% as Other (8.4%; 7.4%). The latter category included participants who identified as American Indian or Alaska Native, or Native Hawaiian or Other Pacific Islander, as well as those who indicated multiple racial identity backgrounds or provided a free-text response for race. Additionally, 11.4% of participants indicated Hispanic ethnicity (12.5%; 9.9%).

The 25^th^, 75^th^ percentiles of HGS were 25 kg and 32 kg for female participants and 37 kg and 50 kg for male participants, respectively. **Table 1** and **Table 2** describe associations between HGS and demographic data according to sex. Younger age was significantly associated with higher HGS in both male and female participants. A plurality (41.8%) of the cohort attended study visits in Palo Alto, CA, while 21.6% were from Kannapolis, NC; 19.7% were from Durham, NC and 16.9% were from Los Angeles, CA. Of the 1375 female participants included after imputation, 300 (21.8%) were ≥65 years of age; 282 (25%) male participants (n=1127) were ≥65 years of age.

**Table 1.** Summary of demographic characteristics for female study participants across hand grip strength classifications. P values calculated using Spearman correlation or Cochran-Armitage tests.

| Demographics | Female Participants by Grip Strength |  |  | P value |
| --- | --- | --- | --- | --- |
|  | Lowest 25%<br>(<24 kg)<br>(n=352) | Middle 50%<br>(24-31 kg)<br>(n=601) | Highest 25%<br>(≥32 kg)<br>(n=365) |  |
| Median age, y (IQR) | 61.6 (43.5-72.2) | 50.9 (36.6-62.1) | 43.0 (32.4-53.2) | <0.001 |
| Race, n (%) |  |  |  |  |
| White | 255 (72.4) | 382 (63.6) | 209 (57.3) | <0.001 |
| Black | 33 (9.4) | 88 (14.6) | 99 (27.1) | <0.001 |
| Asian | 30 (8.5) | 57 (9.5) | 22 (6.0) | 0.22 |
| Other | 30 (8.5) | 74 (12.3) | 35 (9.6) | 0.66 |
| Native Hawaiian/Pacific Islander | 4 (1.1) | 8 (1.3) | 2 (0.5) |  |
| American Indian or Alaska Native | 6 (1.7) | 5 (0.8) | 7 (1.9) |  |
| Other Race with Free-Text Response | 24 (6.8) | 61 (10.1) | 26 (7.1) |  |
| Hispanic ethnicity, n (%) | 50 (14.2) | 75 (12.5) | 40 (11.0) | 0.19 |
| Site, n (%) |  |  |  |  |
| Durham, NC | 44 (12.5) | 112 (18.6) | 122 (33.4) | <0.001 |
| Kannapolis, NC | 112 (31.8) | 119 (19.8) | 64 (17.5) | <0.001 |
| Los Angeles, CA | 39 (11.1) | 128 (21.3) | 62 (17.0) | 0.04 |
| Palo Alto, CA | 157 (44.6) | 242 (40.3) | 117 (32.1) | 0.06 |

**Table 2.** Summary of demographic characteristics for male study participants across hand grip strength classifications. *P* values calculated using Spearman correlation or Cochran-Armitage tests.

| Demographics | Male Participants by Grip Strength |  |  |  |
| --- | --- | --- | --- | --- |
|  | Lowest 25%<br>(<37 kg)<br>(n=244) | Middle 50%<br>(37-49 kg)<br>(n=534) | Highest 25%<br>(≥50 kg)<br>(n=270) | *P* value |
| Median age, y (IQR) | 66.0 (47.1-74.4) | 51.2 (34.3-64.7) | 41.4 (31.1-50.7) | <0.001 |
| Race, n (%) |  |  |  |  |
| White | 160 (65.6) | 349 (65.4) | 147 (54.4) | 0.008 |
| Black | 27 (11.1) | 76 (14.2) | 57 (21.1) | 0.001 |
| Asian | 41 (16.8) | 63 (11.8) | 30 (11.1) | 0.06 |
| Other | 16 (6.6) | 46 (8.6) | 36 (13.3) | 0.008 |
| Native Hawaiian/Pacific Islander | 2 (0.8) | 4 (0.7) | 5 (1.9) |  |
| American Indian or Alaska Native | 0 (0) | 5 (0.9) | 4 (1.5) |  |
| Other Race with Free-Text Response | 14 (5.7) | 37 (6.9) | 27 (10.0) |  |
| Hispanic ethnicity, n (%) | 21 (8.6) | 55 (10.3) | 28 (10.4) | 0.51 |
| Site, n (%) |  |  |  |  |
| Durham, NC | 28 (11.5) | 99 (18.5) | 61 (22.6) | 0.001 |
| Kannapolis, NC | 64 (26.2) | 95 (17.8) | 57 (21.1) | 0.18 |
| Los Angeles, CA | 19 (7.8) | 100 (18.7) | 53 (19.6) | <0.001 |
| Palo Alto, CA | 133 (54.5) | 240 (44.9) | 99 (36.7) | <0.001 |

An association between Black race and higher HGS was seen in all female participants, regardless of age (**Figure 1a** and **Figure 1b**). In regression models using variables selected from the MI-LASSO, (**Table 3**) Black race was associated with approximately 3-4 kg higher HGS in female participants regardless of age group. No other variables identified by MI-LASSO were shared between female age groups. Among female participants, we identified several variables associated with HGS that were specific to age group. Of those identified, the categorical variables associated with ≥2 kg lower HGS among younger female participants were history of breast cancer, osteopenia, and transient ischemic attack (TIA). Continuous variables where higher values were associated with lower HGS in younger female participants included estimated glomerular filtration rate (eGFR), World Health Organization (WHO) Disability Assessment Schedule (WHODAS 2.0) total score, and cigarette pack-years smoked. Among older female participants, additional categorical variables associated with ≥2 kg lower HGS were body image concerns and monocyte count. Continuous variables where higher values were associated with lower HGS in this group included low-density-lipoprotein (LDL) cholesterol, Generalized Anxiety Disorder-7 (GAD-7) score, and absolute monocyte count.

**Figure 1a.**
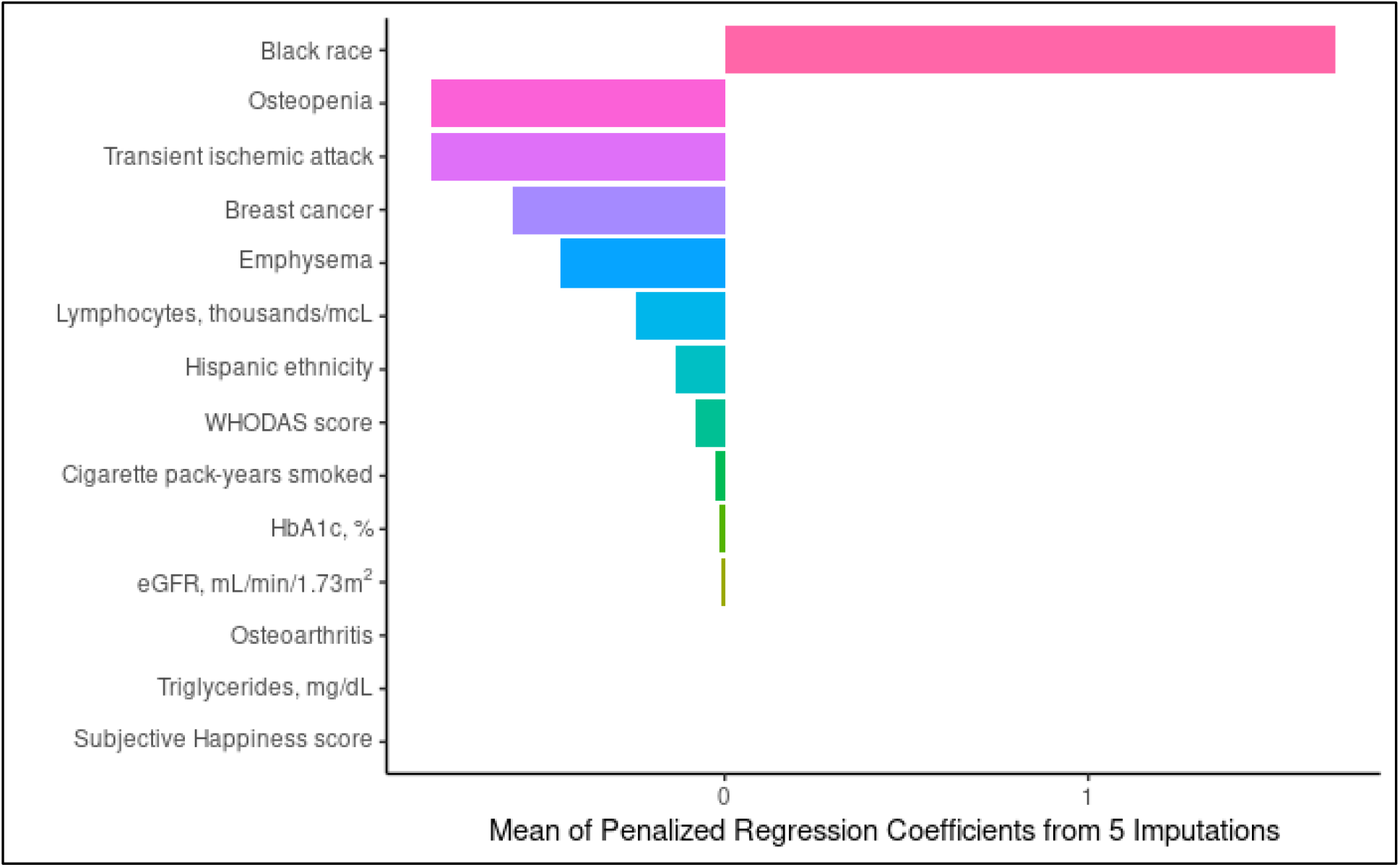
Characteristics associated with hand grip strength after LASSO regression using multiply imputed data: females study participants aged <65 years (N=1075). Abbreviations: WHODAS, World Health Organization Disability Assessment Schedule 2.0; HbA1c, hemoglobin A1c; eGFR, estimated glomerular filtration rate.

**Figure 1b.**
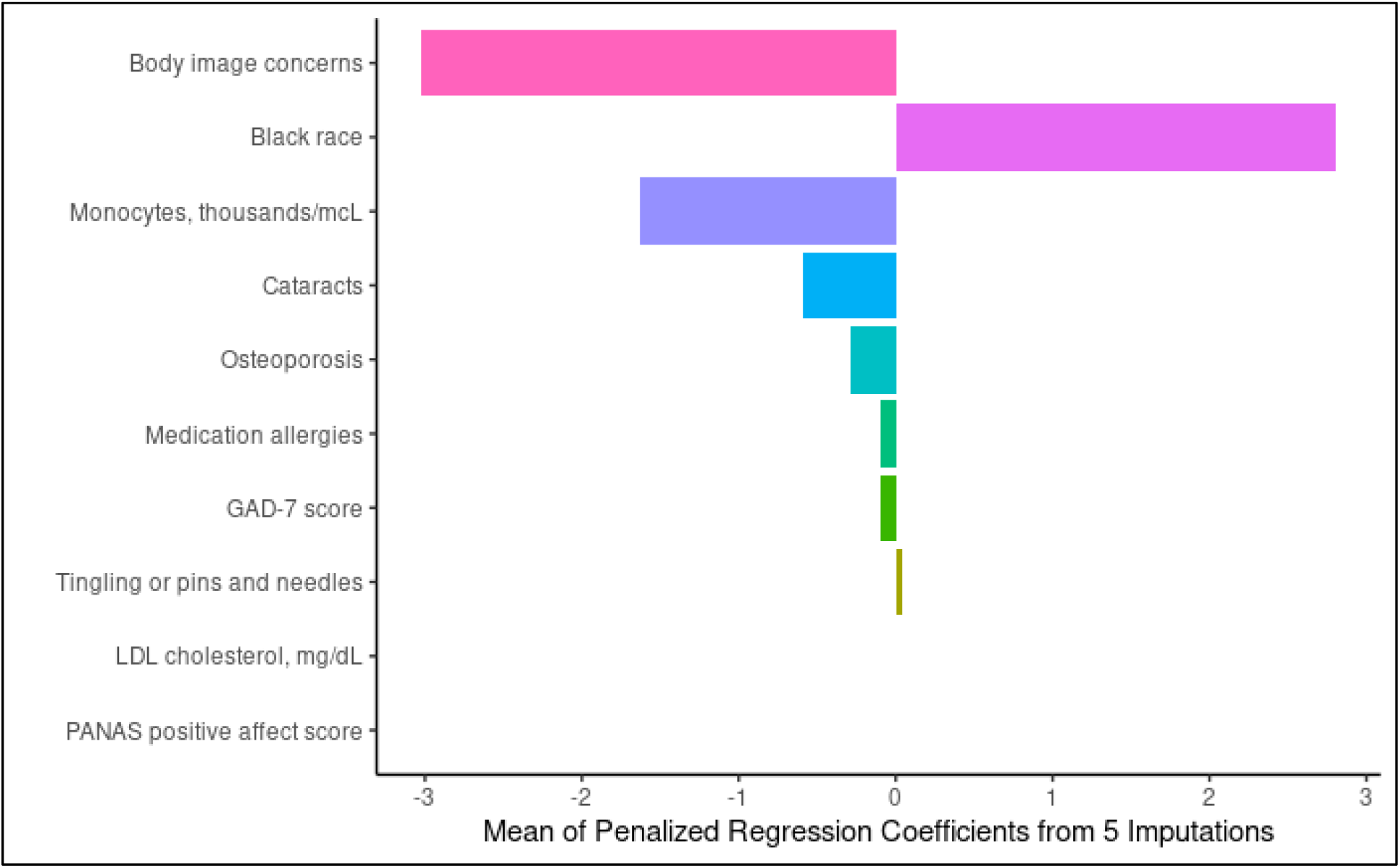
Characteristics Associated with hand grip strength after LASSO regression using multiply imputed data: females study participants ≥65 years (N=300). Because no study participants in this age/sex group had type 1 diabetes mellitus, this variable was removed from the model for this group. Abbreviations: GAD-7, Generalized Anxiety Disorder; LDL, low density lipoprotein; PANAS, Positive and Negative Affect Schedule.

**Figure 1c.**
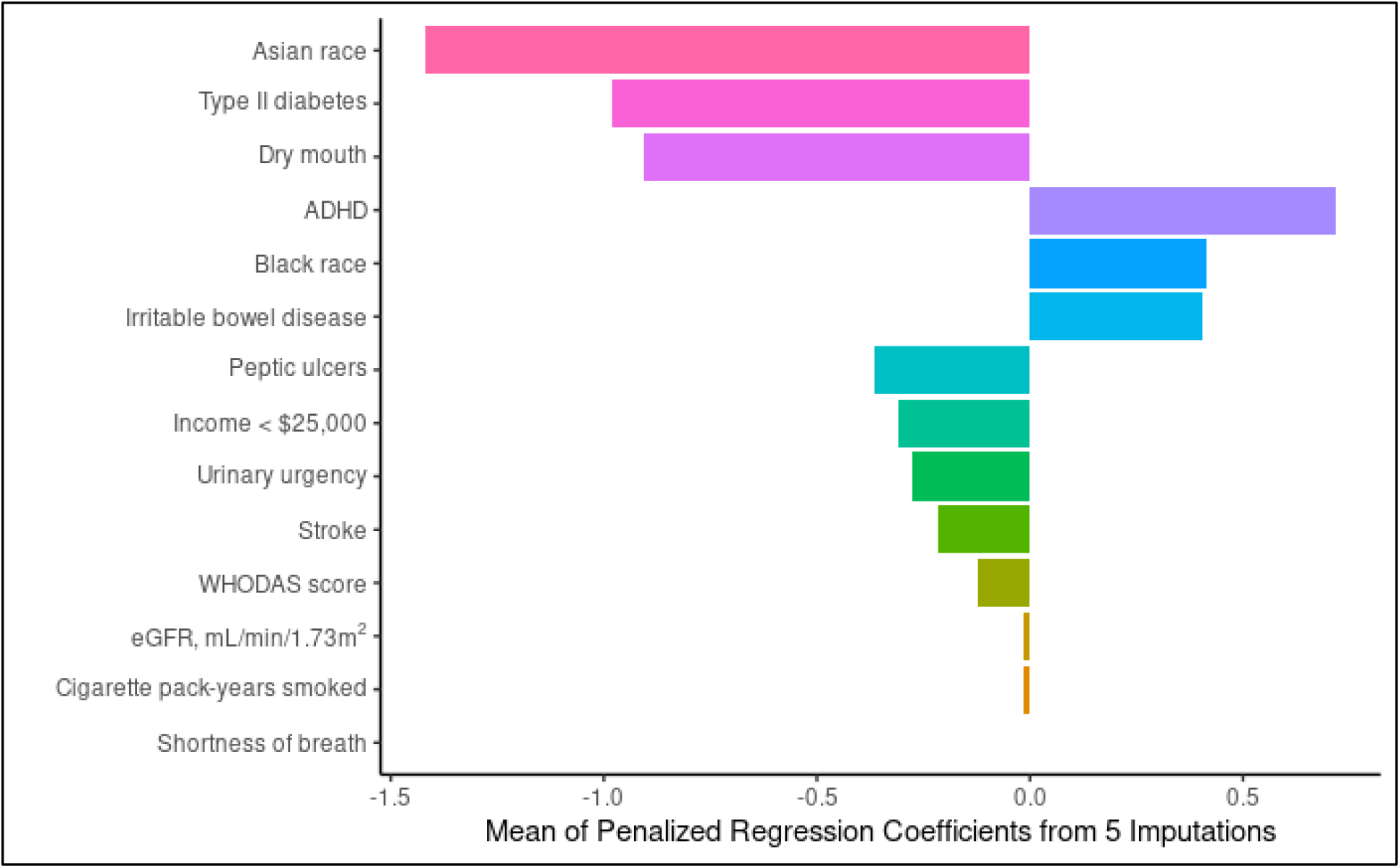
Characteristics associated with hand grip strength after LASSO regression using multiply imputed data: male study participants aged <65 years (N=845). Abbreviations: ADHD, attention-deficit/hyperactivity disorder; WHODAS, World Health Organization Disability Assessment Schedule 2.0, eGFR, estimated glomerular filtration rate.

**Figure 1d.**
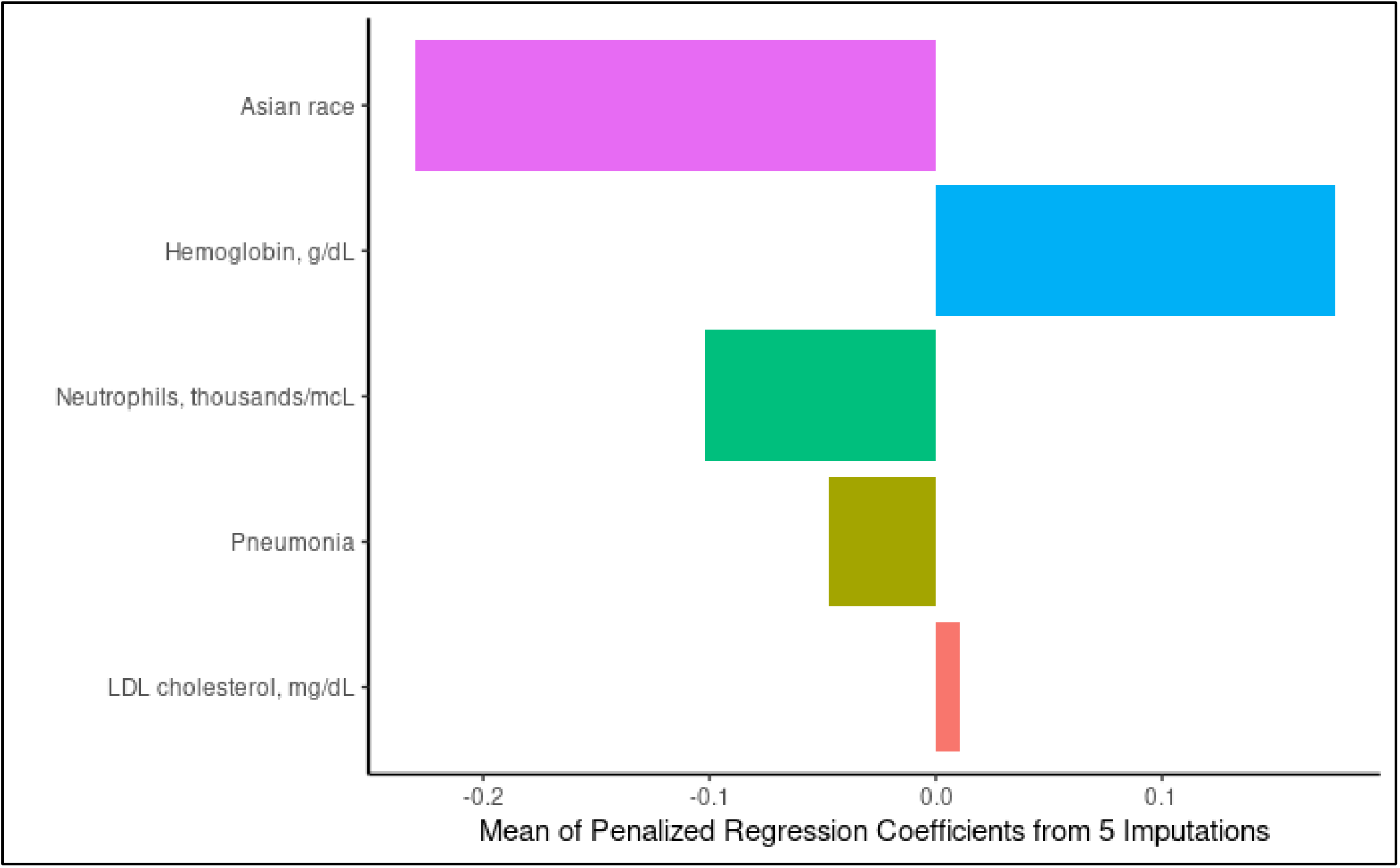
Characteristics associated with hand grip strength after LASSO regression using multiply imputed data: male study participants aged ≥65 years (N=282). Because no study participants in this age/sex group had fibromyalgia, this variable was removed from the model for this group. Abbreviations: LDL, low-density lipoprotein.

**Table 3.**
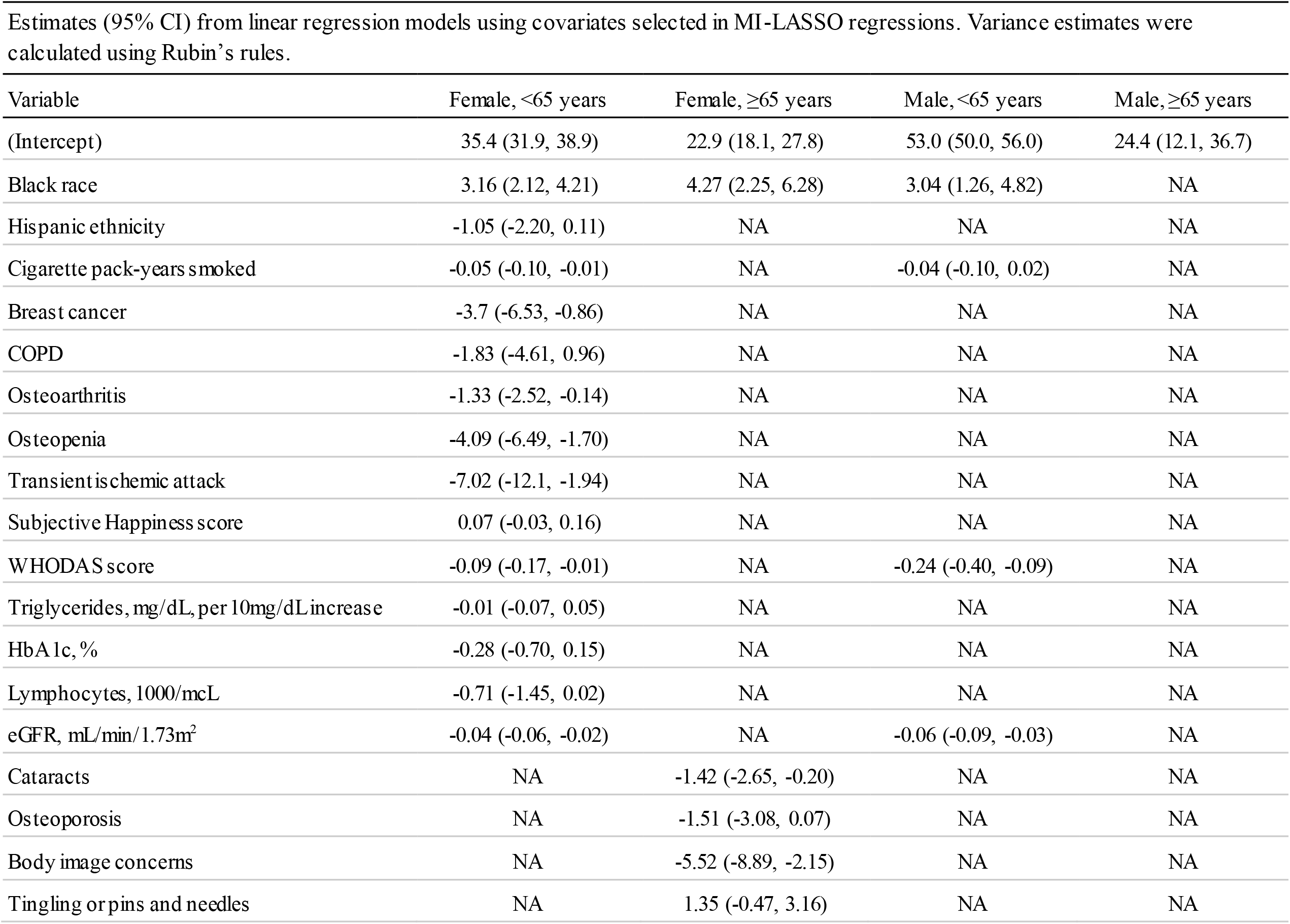
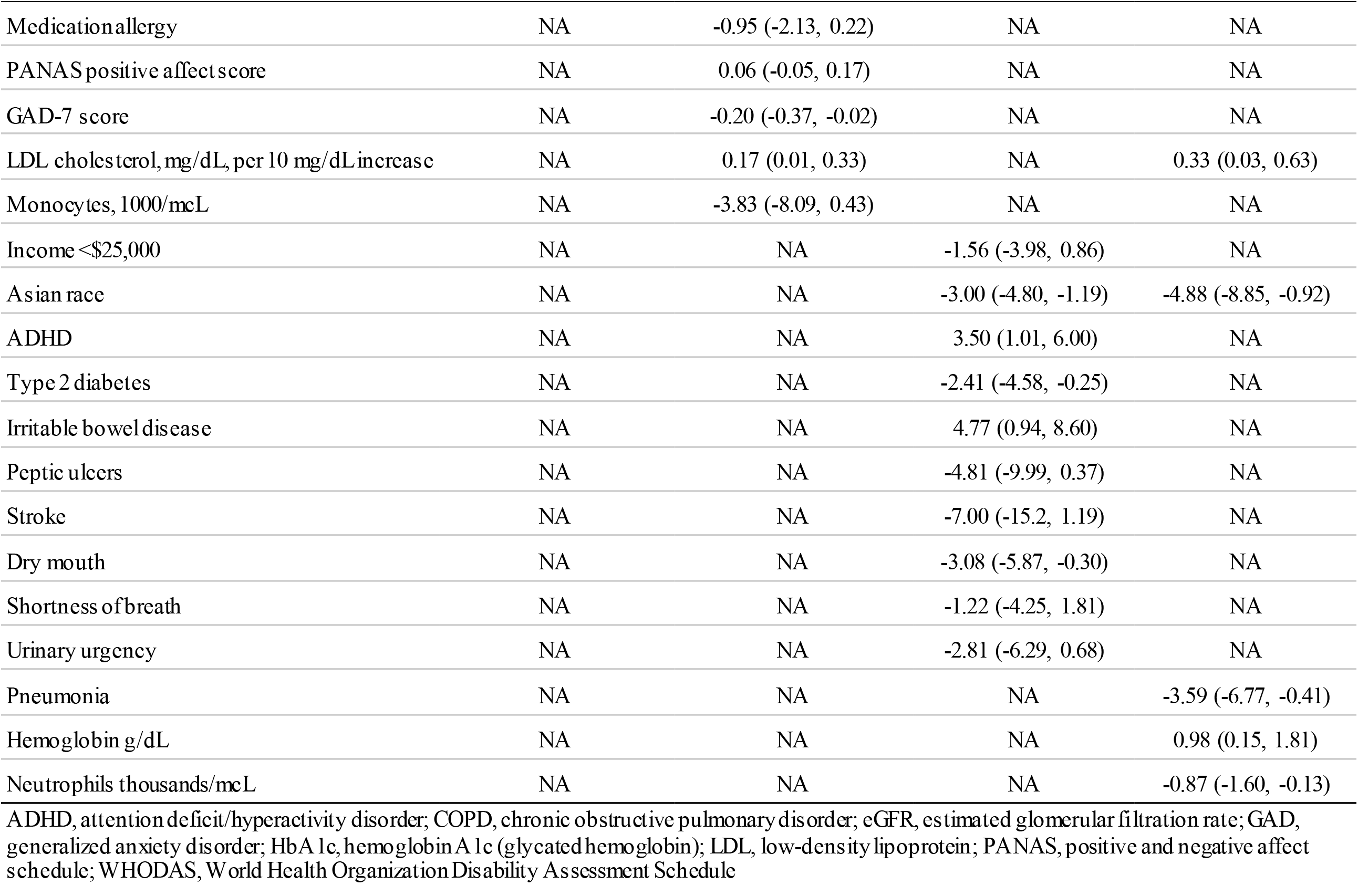
Estimates (95% CI) from linear regression models using covariates selected in MI-LASSO regressions. Variance estimates were calculated using Rubin’s rules.

| Variable | Female, <65 years | Female, ≥65 years | Male, <65 years | Male, ≥65 years |
| --- | --- | --- | --- | --- |
| (Intercept) | 35.4 (31.9, 38.9) | 22.9 (18.1, 27.8) | 53.0 (50.0, 56.0) | 24.4 (12.1, 36.7) |
| Black race | 3.16 (2.12, 4.21) | 4.27 (2.25, 6.28) | 3.04 (1.26, 4.82) | NA |
| Hispanic ethnicity | -1.05 (-2.20, 0.11) | NA | NA | NA |
| Cigarette pack-years smoked | -0.05 (-0.10, -0.01) | NA | -0.04 (-0.10, 0.02) | NA |
| Breast cancer | -3.7 (-6.53, -0.86) | NA | NA | NA |
| COPD | -1.83 (-4.61, 0.96) | NA | NA | NA |
| Osteoarthritis | -1.33 (-2.52, -0.14) | NA | NA | NA |
| Osteopenia | -4.09 (-6.49, -1.70) | NA | NA | NA |
| Transient ischemic attack | -7.02 (-12.1, -1.94) | NA | NA | NA |
| Subjective Happiness score | 0.07 (-0.03, 0.16) | NA | NA | NA |
| WHODAS score | -0.09 (-0.17, -0.01) | NA | -0.24 (-0.40, -0.09) | NA |
| Triglycerides, mg/dL, per 10mg/dL increase | -0.01 (-0.07, 0.05) | NA | NA | NA |
| HbA1c, % | -0.28 (-0.70, 0.15) | NA | NA | NA |
| Lymphocytes, 1000/mcL | -0.71 (-1.45, 0.02) | NA | NA | NA |
| eGFR, mL/min/1.73m<sup>2</sup> | -0.04 (-0.06, -0.02) | NA | -0.06 (-0.09, -0.03) | NA |
| Cataracts | NA | -1.42 (-2.65, -0.20) | NA | NA |
| Osteoporosis | NA | -1.51 (-3.08, 0.07) | NA | NA |
| Body image concerns | NA | -5.52 (-8.89, -2.15) | NA | NA |
| Tingling or pins and needles | NA | 1.35 (-0.47, 3.16) | NA | NA |
| Medication allergy | NA | -0.95 (-2.13, 0.22) | NA | NA |
| PANAS positive affect score | NA | 0.06 (-0.05, 0.17) | NA | NA |
| GAD-7 score | NA | -0.20 (-0.37, -0.02) | NA | NA |
| LDL cholesterol, mg/dL, per 10 mg/dL increase | NA | 0.17 (0.01, 0.33) | NA | 0.33 (0.03, 0.63) |
| Monocytes, 1000/mcL | NA | -3.83 (-8.09, 0.43) | NA | NA |
| Income <\$25,000 | NA | NA | -1.56 (-3.98, 0.86) | NA |
| Asian race | NA | NA | -3.00 (-4.80, -1.19) | -4.88 (-8.85, -0.92) |
| ADHD | NA | NA | 3.50 (1.01, 6.00) | NA |
| Type 2 diabetes | NA | NA | -2.41 (-4.58, -0.25) | NA |
| Irritable bowel disease | NA | NA | 4.77 (0.94, 8.60) | NA |
| Peptic ulcers | NA | NA | -4.81 (-9.99, 0.37) | NA |
| Stroke | NA | NA | -7.00 (-15.2, 1.19) | NA |
| Dry mouth | NA | NA | -3.08 (-5.87, -0.30) | NA |
| Shortness of breath | NA | NA | -1.22 (-4.25, 1.81) | NA |
| Urinary urgency | NA | NA | -2.81 (-6.29, 0.68) | NA |
| Pneumonia | NA | NA | NA | -3.59 (-6.77, -0.41) |
| Hemoglobin g/dL | NA | NA | NA | 0.98 (0.15, 1.81) |
| Neutrophils thousands/mcL | NA | NA | NA | -0.87 (-1.60, -0.13) |

ADHD, attention deficit/hyperactivity disorder; COPD, chronic obstructive pulmonary disorder; eGFR, estimated glomerular filtration rate; GAD, generalized anxiety disorder; HbA 1c, hemoglobin A 1c (glycated hemoglobin); LDL, low-density lipoprotein; PANAS, positive and negative affect schedule; WHODAS, World Health Organization Disability Assessment Schedule

Among male participants (**Figure 1c; Figure 1d; Table 3**), Asian race was associated with approximately 3-5 kg lower HGS; Black race was associated with approximately 3 kg higher HGS among younger male participants. Categorical variables associated with ≥2 kg lower HGS included type 2 diabetes mellitus (T2DM), dry mouth, peptic ulcers, and stroke in younger male participants, while attention-deficit/hyperactivity disorder (ADHD) and irritable bowel disease were associated with ≥2 kg higher HGS. Continuous variables for which higher values were associated with lower HGS in this group included eGFR and WHODAS 2.0 score. In addition to Asian race, pneumonia was associated with ≥2 kg lower HGS in older male participants. Additionally, higher hemoglobin and LDL cholesterol levels were associated with higher HGS while higher neutrophil count was associated with lower HGS in older male participants.

## Discussion

We sought to identify the most salient factors associated with HGS out of a wide range of clinical variables when stratifying by both sex and age as a means of high-throughput hypothesis generation. Our study is the first to stratify by age in order to investigate variables associated with HGS in a large, deeply phenotyped cohort, allowing comparisons across both sex and age. Our results underscore previous findings that HGS is associated with several health dimensions of interest and that these associations vary by age and sex. In addition to confirming a number of previously reported associations, our analyses also identified several novel associations with HGS that warrant further investigation in specific age and sex groups. Novel associations include eGFR in individuals aged <65, LDL cholesterol in individuals aged ≥65, psychosocial factors such as body image concerns, subjective happiness, and PANAS positive affect score in females and ADHD in males aged <65, and immune cells in males aged ≥65 and females overall.

Race was consistently associated with HGS across sex and age categories to varying degrees, findings that are consistent with published literature reporting differences in HGS across race categories [2,32,85,86]. These observed associations may be explained at least in part by differences in body composition that are often seen across differing ethnic or racial backgrounds [87,88]; however, the reasons for these observed differences are likely complex, as social factors and experiences associated with race such as health and wealth disparities, impacts of racism, and other unmeasured racialized experiences will be jointly captured by the race variable in our regression models [89].

The only other factor commonly associated with HGS across more than 2 stratified groups was immune cell count; however, the specific type of white blood cell associated with HGS differed between groups. An association between higher absolute monocyte count and lower HGS was observed in female participants aged ≥65 years, while an association between higher total lymphocyte count and lower HGS was seen in female participants aged <65 years. Among male participants aged ≥65 years, higher total neutrophil count was also associated with lower HGS, but no measure of immune cells was associated with HGS among younger male participants. Few studies have reported on associations between HGS and immune cells, although one reported an association between neutrophil dysfunction and age that was exaggerated by frailty (for which HGS is an indicator) in older adults [90]. Additionally, only one study has reported on the association between heat shock protein expression in monocytes and lymphocytes with HGS in older adults [91]. In our study, absolute monocyte count was associated with the largest estimated decrease in predicted HGS, with each 1-unit increase in absolute monocyte count being associated with a nearly 4-kg lower HGS. Immune cells other than absolute monocyte count were associated with HGS in the other groups to a lesser degree, with the estimated HGS being <1 kg lower per 1-unit increase.

In addition to sharing associations between race and HGS, female participants in our study also shared associations between bone mineral density conditions and HGS across age groups. Previous studies have reported an association between low HGS in older adults (primarily perimenopausal and postmenopausal women) and insufficient bone mineral density at sites remote from the upper extremity (e.g., spine and hip) [92-100]. In addition, HGS has been identified as a viable supplemental predictor of incident fractures when traditional bone mineral density scans (i.e., dual-energy x-ray absorptiometry) are unavailable or cost-prohibitive [77-82]. Our results, which show that lower HGS is associated with osteoporosis in older female participants and with osteopenia in female participants aged <65 years, provide additional support for these reported associations between HGS and bone mineral density. Although we did not observe associations between HGS and prevalent osteopenia or osteoporosis in male participants, others have reported this association in a large, nationally representative sample when limiting their analysis to adults aged ≥40 years [101].

Other than race and bone mineral density conditions, no variables associated with HGS were common across age groups among female participants. In female participants aged <65, several of the diagnoses associated with lower HGS are diagnoses associated with worse physical function and overall strength: breast cancer, TIA, COPD, and osteoarthritis.

Younger female participants showed a small association between higher subjective happiness scores and higher HGS; however, female participants aged ≥65 had multiple indicators of mental health associated with HGS. Most notable among these were body image concerns and GAD-7 score, both of which were associated with lower HGS. An association between body image concerns and lower HGS in older adults was also reported recently in a study using data from the Korea National Health and Nutrition Examination Survey. In this study, the authors reported that HGS was associated with accuracy of weight estimation and behaviors related to weight loss or maintenance in both male and female participants aged ≥60 years, but not among younger study participants [102].

A number of studies have investigated associations between depression and HGS; however, only 2 report associations between prevalent GAD and HGS. The first, an Irish study in adults aged ≥50 years, found an association between prevalent GAD and HGS in both female and male participants [103]. The second study reported lower HGS in female participants with GAD compared with male participants, especially with those with age of onset ≥40 years [104].

We observed no common factors associated with HGS across age categories other than race, but each age group did have different factors associated with HGS not yet reported. In male study participants aged ≥65, higher hemoglobin levels were associated with higher HGS, while a history of pneumonia was associated with lower HGS. In male participants aged <65 years, a history of ADHD or irritable bowel disease was associated with higher HGS. Although it has been previously demonstrated that ADHD in children has been associated with multiple domains of upper limb function [105], our study is the first to report an association between ADHD and HGS in adults. Similar to younger female participants, younger male participants had some diagnoses associated with lower HGS that are generally associated with worse physical function and overall strength, such as T2DM and stroke.

Symptoms unexpectedly associated with lower HGS were dry mouth (HGS approximately 3 kg lower) and peptic ulcers (approximately 5 kg lower). To our knowledge, associations between these variables and HGS have not been previously reported. However, we note that the association between HGS and dry mouth may be due at least in part to medication data not being available for inclusion in our analyses. This younger male group was also the only group overall to also have a socioeconomic status indicator associated with HGS, with income <$25,000 associated with lower HGS.

Overall, more variables were associated with HGS in participants aged <65 years regardless of sex. In this age group, higher WHODAS 2.0 score, pack-years of smoking, and eGFR were associated with lower HGS. The WHODAS 2.0, a standardized generic measure of health and disability, comprises 6 domains relating to cognition, mobility, self-care, getting along, life activities, and participation [106]. Our finding that a higher total WHODAS 2.0 score (i.e., worse disability) was associated with lower HGS is not unexpected, given previously reported associations between HGS and disability domains either directly (e.g., cognition, mobility, physical function, activity) [2-6,30,31,67,68] or indirectly (e.g., symptoms associated with depression) [24-29] measured by WHODAS 2.0 where these domains have been ascertained using other tools. However, studies linking WHODAS 2.0 disability domains (using alternative metrics) and HGS were done primarily in older adult cohorts, and WHODAS 2.0 score was not associated with HGS for those aged ≥65 years in our study. Although both female and male participants <65 demonstrated this association, the decrease in estimated HGS per unit increase (worse function) in WHODAS 2.0 was more than double for male participants.

Smoking status has likewise been associated with HGS and other measures of physical function in older adults, an association not observed in older adults in this study [107,108]. This association has also been studied in a cohort of British participants aged <65, but although the study reported an association between pack-years of smoking and other physical function measures, there was no association with HGS [109]. However, in our study this association was relatively weak, requiring a 20-pack-year smoking history to be associated with an approximate 1-kg reduction in HGS.

Higher eGFR was also associated with lower HGS in participants aged <65 years but was not associated with HGS in participants ≥65. These findings are consistent with previous studies showing that eGFR, when calculated according to the method used in this study, is not associated with HGS in older adults [110,111]. However, alternative methods for calculating eGFR have been associated with HGS in older adults in at least one other study [112]. Findings in our younger age group indicate that higher eGFR (better kidney function) is associated with lower HGS. The apparent paradox of better kidney function being associated with lower HGS may, like similar paradoxes [113,114], be due to collider bias, which can be induced when selection into the study is affected by both the variable of interest (eGFR) and the outcome (HGS), or when some other third variable affected by these two variables is adjusted for in statistical analysis [115,116]. To our knowledge, no other studies have examined the association between eGFR and HGS in participants aged <65 years.

Among study participants aged ≥65 years, higher LDL cholesterol level was associated with higher HGS, regardless of sex. Although this association was present for both older female and male participant groups, the estimated increase in HGS per unit increase in LDL was twice as high in males. Previously reported associations between LDL and HGS from older adult cohorts in other countries are consistent with this paradoxical association between higher HGS and higher levels of LDL [117,118]. Previous studies have also linked HGS with cardiovascular disease and mortality [51,53-55] and HGS with other indicators of cardiometabolic health/risk (e.g., high-density-lipoprotein cholesterol; blood pressure) [117,119]. However, LDL was the only cardiometabolic risk factor in our results associated with HGS in older adults of either sex.

We note a number of limitations to our study. First, we used cross-sectional data and methods focused on identifying factors most strongly associated with HGS as a means of high-throughput hypothesis generation rather than attempting to estimate the effect of a single specific variable on HGS that would necessitate an alternative approach [120]. As a result, some of the associations described (e.g., eGFR, LDL) may seem paradoxical if interpreted under the assumption that they reflect a causal mechanism. Second, our study sample was limited to populations in North Carolina (Durham and Kannapolis) and California (Los Angeles, Palo Alto). The sample comprised adults older than the national average and had less Hispanic representation (11.4%) than the overall U.S. population (18.5%) [121]. Because of this, the generalizability of our findings to the overall U.S. adult population may be limited.

This study identified several novel salient factors associated with HGS when stratifying by sex and age (<65 versus ≥65 years) and identified some factors that confirm previously published work. Some of these prevalent factors were common across age groups or sex groups, but several factors associated with HGS were specific to both age and sex. These results support the hypothesis that HGS may be a useful indicator of a variety of prevalent conditions, as it was associated with a range of clinical conditions, complaints, and laboratory blood measures. Novel associations identified may merit further investigation into their associations with HGS. If our novel associations are confirmed, then HGS may also be a useful indicator of these variables and further research would be warranted to estimate the direction and magnitude of any causal relationships between HGS and novel factors where biologically plausible.

## Methods

### Project Baseline Health Study (PBHS)

The PBHS (ClinicalTrials.gov identifier NCT03154346) is an ongoing longitudinal multicenter observational cohort study of U.S. adults. Participants contribute demographic, clinical, imaging, and laboratory data at annual visits. The PBHS is funded by Verily Life Sciences and managed in collaboration with Stanford University, Duke University, and the California Health and Longevity Institute. Written informed consent is obtained from all participants enrolled in the PBHS and the study is approved by both a central institutional review board (IRB) (Western IRB) and individual IRBs at each of the participating institutions. All study procedures and study data collection are conducted in accordance with the principles described in the Declaration of Helsinki and with relevant local, state, national, and international standards. Detailed descriptions of study procedures and methods, including exclusion and inclusion criteria and institutional review board approval and enrollment procedures, have been published [83,84].

In this study, we examined cross-sectional data collected at study participants’ baseline visit or collected remotely at that time via survey or connected device. Participants were enrolled through sites affiliated with Stanford University (Palo Alto, CA), Duke University (Durham, NC and Kannapolis, NC), and California Health and Longevity Institute (CHLI; Los Angeles, CA).

### Hand grip strength

HGS was recommended to be measured using the Jamar Plus+ Hand Dynamometer (Patterson Medical). Participants were asked to perform 3 trials measuring peak grip force (kg) with a 10-20 second break between each trial for dominant and non-dominant hands. If the difference between any 2 measures on a single hand was greater than 2.99 kg, a fourth trial was done after a 1-minute resting period. For the purposes of this analysis, HGS was defined as the mean of the dominant hand from the 3-4 trials.

### Other variables

A detailed description of data collected from BHS participants has been previously described [83]. We present a summary of the ascertainment and definition of the 175 variables eligible for entry into each model in the **Appendix**.

### Statistical analysis

Demographic and clinical characteristics including medical conditions, symptoms, patient-reported outcomes, and laboratory measures were described among participants who completed HGS assessments at their baseline visits. Male and female participants were described separately and they were classified into one of 3 cohort-based HGS percentile groups: lowest 25%, middle 50%, and highest 25%.

The Cochran-Armitage test for trend was used to test for associations between HGS and categorical variables of interest, the Spearman rank correlation test was used for continuous variables. Data collected within 200 days of participants’ baseline visits were considered nonmissing. Multiple imputation by chained equations (MICE) methods were used to address missing data [122].

### LASSO regression

Least absolute shrinkage and selection operator procedures addressing multiply imputed data by using group methods [123] (MI-LASSO) were employed to identify variables potentially associated with HGS, which was treated as a continuous outcome. Analyses were performed separately for male and female participants, further stratified by age (<65 years vs. ≥ 65 years), resulting in 4 LASSO regression models. All variables were standardized to allow for meaningful comparison of coefficients. For each age-sex category, shrinkage of coefficients was controlled by a tuning parameter determined by the model that minimized the Bayesian Information Criterion.

Linear regressions using variables selected from the LASSO were performed for each of the 4 age-sex categories to allow for more interpretable coefficient estimates for each variable identified by MI-LASSO. Regression estimates from each imputed dataset were pooled into a single value whose variance was then calculated using Rubin’s rules [124,125].

Analyses were completed using R v3.6.3 [126]. Data were imputed and linear regressions were performed using the MICE package v3.13.0 [122]. LASSO regressions were carried out using the MI.LASSO function [123]. Missing data were imputed using observational data over a series of 5 instances, with 5 iterations per imputation. Tables were created using Python v3.7.10; figures were created with ggplot2 v3.3.0 [127].

## Supporting information

Supplemental Appendix: List of Candidate Covariates for Multivariable LASSO Regression

## Data Availability

The de-identified Project Baseline Health Study (PBHS) data corresponding to this study are available upon request for the purpose of examining its reproducibility. Interested investigators should direct requests to. Requests are subject to approval by PBHS governance.

## Acknowledgments

*Baseline Health Study Team:* American Society of Clinical Oncology, Alexandria, VA, USA: Richard L. Schilsky. Duke University, School of Medicine, Durham, NC, USA: Jennifer Allen, MaryAnn Anderson, Kevin Anstrom, Lucus Araujo, Kristine Arges, Kaveh Ardalan, Bridget Baldwin, Suresh Balu, Mustafa R. Bashir, Manju Bhapkar, Robert Bigelow, Tanya Black, Rosalia Blanco, Gerald Bloomfield, Durga Borkar, Leah Bouk, Ebony Boulware, Nikki Brugnoni, Erin Campbell, Paul Campbell, Larry Carin, Tammy Jo Cassella, Tina Cates, Ranee Chatterjee Montgomery, Victoria Christian, John Choong, Michael Cohen-Wolkowiez, Elizabeth Cook, Scott Cousins, Ashley Crawford, Nisha Datta, Melissa Daubert, James Davis, Jillian Dirkes, Isabelle Doan, Marie Dockery, P. Murali Doraiswamy, Pamela S. Douglas, Shelly Duckworth, Ashley Dunham, Gary Dunn, Ryan Ebersohl, Julie Eckstrand, Vivienne Fang, April Flora, Emily Ford, Lucia Foster, Elizabeth Fraulo, John French, Geoffrey S. Ginsburg, Cindy Green, Latoya Greene, Jeffrey Guptill, Donna Hamel, Jennifer Hamill, Chris Harrington, Rob Harrison, Lauren Hedges, Brooke Heidenfelder, Adrian F. Hernandez, Cindy Heydary, Tim Hicks, Lina Hight, Deborah Hopkins, Erich S. Huang, Grace Huh, Jillian Hurst, Kelly Inman, Gemini Janas, Glenn Jaffee, Janace Johnson, Tiffanie Keaton, Michel Khouri, Daniel King, Jennifer Korzekwinski, Lynne H. Koweek, Anthony Kuo, Lydia Kwee, Dawn Landis, Rachele Lipsky, Desiree Lopez, Carolyn Lowry, Kelly Marcom, Keith Marsolo, Paige McAdams, Shannon McCall, Robert McGarrah, John McGugan, Dani Mee, Sabrena Mervin-Blake, Prithu Mettu, Mathias Meyer, Justin Meyers, Calire N. Miller, Rebecca Moen, Lawrence H. Muhlbaier, Michael Murphy, Ben Neely, L. Kristin Newby, Jayne Nicoldson, Hoang Nguyen, Maggie Nguyen, Lori O’Brien, Sumru Onal, Jeremey O’Quinn, David Page, Neha J. Pagidipati, Kishan Parikh, Sarah R. Palmer, Bray Patrick-Lake, Brenda Pattison, Michael Pencina, Eric D. Peterson, Jon Piccini, Terry Poole, Tom Povsic, Alicia Provencher, Dawn Rabineau, Annette Rich, Susan Rimmer, Fides Schwartz, Angela Serafin, Nishant Shah, Svati Shah, Kelly Shields, Steven Shipes, Peter Shrader, Jon Stiber, Lynn Sutton, Geeta Swamy, Betsy Thomas, Sandra Torres, Debara Tucci, Anthony Twisdale, Susan A. Whitney, Robin Williamson, Lauren Wilverding, Charlene A. Wong, Lisa Wruck. Ellen Young Gemini Group, USA: Jane Perlmutter. Health Collaboratory and Cancer 101, New York, NY, USA: Sarah Krug. Rare Dots, Inc., USA: S. Whitney Bowman-Zatzkin. Society of Participatory Medicine, USA: Sarah Krug. Stanford University, School of Medicine, Stanford, CA, USA: Themistocles Assimes, Vikram Bajaj, Maxwell Cheong, Millie Das, Manisha Desai, Alice C. Fan, Dominik Fleischmann, Sanjiv S. Gambhir, Garry Gold, Francois Haddad, David Hong, Curtis Langlotz, Yaping J. Liao, Rong Lu, Kenneth W. Mahaffey, David Maron, Rebecca McCue, Rajan Munshi, Fatima Rodriguez, Sumana Shashidhar, George Sledge, Susie Spielman, Ryan Spitler, Sue Swope, Donna Williams, Julio C. Nunes. University of Florida, College of Medicine, Gainesville, FL, USA: Carl J Pepine. University of Missouri, Children’s Mercy Hospital, Kansas City, MO, USA: John D Lantos. University of Texas, Dell Medical School, Austin, TX, USA: Michael Pignone. University of Washington, Department of Biostatistics, Seattle, WA, USA: Patrick Heagerty. Vanderbilt University, School of Medicine, Nashville, TN, USA: Laura Beskow, Gordon Bernard. Verily Inc., South San Francisco, CA, USA: Kelley Abad, Giulia Angi, Robert M. Califf, Lawrence Deang, Joy Huynh, Manway Liu, Cherry Mao, Michael Magdaleno, William J. Marks, Jr., Jessica Mega, David Miller, Nicole Ong, Darshita Patel, Vanessa Ridaura, Scarlet Shore, Sarah Short, Michelle Tran, Veronica Vu, Celeste Wong. Harvard University, School of Medicine, Boston, MA, USA: Robert C. Green. Google Inc., Mountain View, CA, USA: John Hernandez. California Health and Longevity Institute, Westlake Village, CA, USA: Jolene Benge, Gislia Negrete, Gelsey Sierra, Terry Schaack

## Author contributions

Kenneth A. Taylor – Conceptualization, Methodology, Writing – Original draft preparation, Project administration

Megan K. Carroll - Conceptualization, Methodology, Formal analysis, Visualization, Writing – Original draft preparation

Sarah Short – Methodology, Data curation, Writing – Review and Editing Adam P. Goode – Writing – Review and Editing

## References

1. Sanderson, W. C., Scherbov, S., Weber, D. & Bordone, V. Combined measures of upper and lower body strength and subgroup differences in subsequent survival among the older population of England. J. Aging Health 28, 1178–1193 (2016).

2. Forrest, K. Y. Z., Williams, A. M., Leeds, M. J., Robare, J. F. & Bechard, T. J. Patterns and correlates of grip strength in older Americans. Curr. Aging Sci. 11, 63–70 (2018).

3. Wang, C.-Y. & Chen, L.-Y. Grip strength in older adults: test-retest reliability and cutoff for subjective weakness of using the hands in heavy tasks. Arch. Phys. Med. Rehabil. 91, 1747–1751 (2010).

4. Zhang, Q. et al. 6MWT Performance and its Correlations with VO_2_ and handgrip strength in home-dwelling mid-aged and older Chinese. Int. J. Environ. Res. Public Health 14, (2017).

5. Vasconcelos, K. S. de S. et al. Handgrip strength cutoff points to identify mobility limitation in community-dwelling older people and associated factors. J. Nutr. Health Aging 20, 306–315 (2016).

6. Sallinen, J. et al. Hand-grip strength cut points to screen older persons at risk for mobility limitation. J. Am. Geriatr. Soc. 58, 1721–1726 (2010).

7. Hu, S. et al. Relationship between grip strength and prediabetes in a large-scale adult population. Am. J. Prev. Med. 56, 844–851 (2019).

8. Andersen, H., Nielsen, S., Mogensen, C. E. & Jakobsen, J. Muscle strength in type 2 diabetes. Diabetes 53, 1543–1548 (2004).

9. Bohannon, R. W., Smith, J. & Barnhard, R. Grip strength in end stage renal disease. Percept. Mot. Skills 79, 1523–1526 (1994).

10. Lee, M.-R., Jung, S. M., Bang, H., Kim, H. S. & Kim, Y. B. Association between muscle strength and type 2 diabetes mellitus in adults in Korea: Data from the Korea national health and nutrition examination survey (KNHANES) VI. Medicine 97, e10984 (2018).

11. Peterson, M. D. et al. Low normalized grip strength is a biomarker for cardiometabolic disease and physical disabilities among U.S. and Chinese adults. J. Gerontol. A Biol. Sci. Med. Sci. 72, 1525–1531 (2017).

12. Cheung, C.-L., Nguyen, U.-S. D. T., Au, E., Tan, K. C. B. & Kung, A. W. C. Association of handgrip strength with chronic diseases and multimorbidity: a cross-sectional study. Age 35, 929–941 (2013).

13. Reeve, T. E., IV et al. Grip strength measurement for frailty assessment in patients with vascular disease and associations with comorbidity, cardiac risk, and sarcopenia. J. Vasc. Surg. 67, 1512–1520 (2018).

14. Volaklis, K. A. et al. Handgrip strength is inversely and independently associated with multimorbidity among older women: Results from the KORA-Age study. Eur. J. Intern. Med. 31, 35–40 (2016).

15. Yorke, A. M., Curtis, A. B., Shoemaker, M. & Vangsnes, E. The impact of multimorbidity on grip strength in adults age 50 and older: Data from the health and retirement survey (HRS). Arch. Gerontol. Geriatr. 72, 164–168 (2017).

16. Byrnes, A., Mudge, A., Young, A., Banks, M. & Bauer, J. Use of hand grip strength in nutrition risk screening of older patients admitted to general surgical wards. Nutr. Diet. 75, 520–526 (2018).

17. McNicholl, T. et al. Handgrip strength, but not 5-meter walk, adds value to a clinical nutrition assessment. Nutr. Clin. Pract. 34, 428–435 (2019).

18. Silva, L. F. et al. Handgrip strength as a simple indicator of possible malnutrition and inflammation in men and women on maintenance hemodialysis. J. Ren. Nutr. 21, 235–245 (2011).

19. Zhang, X. S. et al. Handgrip strength as a predictor of nutritional status in Chinese elderly inpatients at hospital admission. Biomed. Environ. Sci. 30, 802–810 (2017).

20. Chen, H.-C., Hsu, N.-W. & Chou, P. The association between sleep duration and hand grip strength in community-dwelling older adults: The Yilan study, Taiwan. Sleep 40, (2017).

21. Laredo-Aguilera, J. A., Carmona-Torres, J. M., Cobo-Cuenca, A. I., García-Pinillos, F. & Latorre-Román, P. Á. Handgrip strength is associated with psychological functioning, mood and sleep in women over 65 years. Int. J. Environ. Res. Public Health 16, (2019).

22. Lee, G., Baek, S., Park, H.-W. & Kang, E. K. Sleep quality and attention may correlate with hand grip strength: FARM Study. Ann. Rehabil. Med. 42, 822–832 (2018).

23. Pengpid, S. & Peltzer, K. Hand grip strength and its sociodemographic and health correlates among older adult men and women (50 Years and Older) in Indonesia. Curr. Gerontol. Geriatr. Res. 2018, 3265041 (2018).

24. Ashdown-Franks, G. et al. Handgrip strength and depression among 34,129 adults aged 50 years and older in six low- and middle-income countries. J. Affect. Disord. 243, 448–454 (2019).

25. Gopinath, B., Kifley, A., Liew, G. & Mitchell, P. Handgrip strength and its association with functional independence, depressive symptoms and quality of life in older adults. Maturitas 106, 92–94 (2017).

26. Han, K.-M. et al. Relationships between hand-grip strength, socioeconomic status, and depressive symptoms in community-dwelling older adults. J. Affect. Disord. 252, 263–270 (2019).

27. Lee, M.-R., Jung, S. M., Bang, H., Kim, H. S. & Kim, Y. B. The association between muscular strength and depression in Korean adults: a cross-sectional analysis of the sixth Korea National Health and Nutrition Examination Survey (KNHANES VI) 2014. BMC Public Health 18, 1123 (2018).

28. Szlejf, C. et al. Depression is associated with sarcopenia due to low muscle strength: Results from the ELSA-Brasil Study. J. Am. Med. Dir. Assoc. 20, 1641–1646 (2019).

29. Gu, Y. et al. Grip strength and depressive symptoms in a large-scale adult population: The TCLSIH cohort study. J. Affect. Disord. 279, 222–228 (2021).

30. Vancampfort, D. et al. Associations between handgrip strength and mild cognitive impairment in middle-aged and older adults in six low- and middle-income countries. Int. J. Geriatr. Psychiatry 34, 609–616 (2019).

31. Kobayashi-Cuya, K. E. et al. Observational evidence of the association between handgrip strength, hand dexterity, and cognitive performance in community-dwelling older adults: A systematic review. J. Epidemiol. 28, 373–381 (2018).

32. Wang, Y.-C., Bohannon, R. W., Li, X., Sindhu, B. & Kapellusch, J. Hand-grip strength: normative reference values and equations for individuals 18 to 85 years of age residing in the United States. J. Orthop. Sports Phys. Ther. 48, 685–693 (2018).

33. Wong, S. L. Grip strength reference values for Canadians aged 6 to 79: Canadian Health Measures Survey, 2007 to 2013. Health Rep. 27, 3–10 (2016).

34. Peterson, M. D. & Krishnan, C. Growth charts for muscular strength capacity with quantile regression. Am. J. Prev. Med. 49, 935–938 (2015).

35. Dodds, R. M. et al. Grip strength across the life course: normative data from twelve British studies. PLoS One 9, e113637 (2014).

36. Massy-Westropp, N. M., Gill, T. K., Taylor, A. W., Bohannon, R. W. & Hill, C. L. Hand grip strength: age and gender stratified normative data in a population-based study. BMC Res. Notes 4, 127 (2011).

37. Schlüssel, M. M., dos Anjos, L. A., de Vasconcellos, M. T. L. & Kac, G. Reference values of handgrip dynamometry of healthy adults: a population-based study. Clin. Nutr. 27, 601–607 (2008).

38. Bohannon, R. W., Peolsson, A., Massy-Westropp, N., Desrosiers, J. & Bear-Lehman, J. Reference values for adult grip strength measured with a Jamar dynamometer: a descriptive meta-analysis. Physiotherapy 92, 11–15 (2006).

39. Adedoyin, R. A. et al. Reference values for handgrip strength among healthy adults in Nigeria. Hong Kong Physiother. J. 27, 21–29 (2009).

40. Frederiksen, H. et al. Age trajectories of grip strength: cross-sectional and longitudinal data among 8,342 Danes aged 46 to 102. Ann. Epidemiol. 16, 554–562 (2006).

41. Spruit, M. A., Sillen, M. J. H., Groenen, M. T. J., Wouters, E. F. M. & Franssen, F. M. E. New normative values for handgrip strength: results from the UK Biobank. J. Am. Med. Dir. Assoc. 14, 775.e5–11 (2013).

42. Alley, D. E. et al. Grip strength cutpoints for the identification of clinically relevant weakness. J. Gerontol. A Biol. Sci. Med. Sci. 69, 559–566 (2014).

43. Duchowny, K. Do nationally representative cutpoints for clinical muscle weakness predict mortality? Results from 9 years of follow-up in the Health and Retirement Study. J. Gerontol. A Biol. Sci. Med. Sci. 74, 1070–1075 (2019).

44. Duchowny, K. A., Peterson, M. D. & Clarke, P. J. Cut points for clinical muscle weakness among older Americans. Am. J. Prev. Med. 53, 63–69 (2017).

45. Bohannon, R. W. Grip strength: an indispensable biomarker for older adults. Clin. Interv. Aging 14, 1681–1691 (2019).

46. Bahat, G. et al. Cut-off points to identify sarcopenia according to European Working Group on Sarcopenia in Older People (EWGSOP) definition. Clin. Nutr. 35, 1557–1563 (2016).

47. de Souza Barbosa, J. F. et al. Clinically relevant weakness in diverse populations of older adults participating in the International Mobility in Aging Study. Age 38, 25 (2016).

48. Studenski, S. A. et al. The FNIH sarcopenia project: rationale, study description, conference recommendations, and final estimates. J. Gerontol. A Biol. Sci. Med. Sci. 69, 547–558 (2014).

49. Dodds, R. M. et al. Global variation in grip strength: a systematic review and meta-analysis of normative data. Age Ageing 45, 209–216 (2016).

50. Roberts, H. C. et al. A review of the measurement of grip strength in clinical and epidemiological studies: towards a standardised approach. Age Ageing 40, 423–429 (2011).

51. Wu, Y., Wang, W., Liu, T. & Zhang, D. Association of grip strength with risk of all-cause mortality, cardiovascular diseases, and cancer in community-dwelling populations: a meta-analysis of prospective cohort studies. J. Am. Med. Dir. Assoc. 18, 551.e17–551.e35 (2017).

52. Gubelmann, C., Vollenweider, P. & Marques-Vidal, P. No association between grip strength and cardiovascular risk: The CoLaus population-based study. Int. J. Cardiol. 236, 478–482 (2017).

53. Prasitsiriphon, O. & Pothisiri, W. Associations of grip strength and change in grip strength with all-cause and cardiovascular mortality in a European older population. Clin. Med. Insights Cardiol. 12, 1179546818771894 (2018).

54. Yates, T. et al. Association of walking pace and handgrip strength with all-cause, cardiovascular, and cancer mortality: a UK Biobank observational study. Eur. Heart J. 38, 3232–3240 (2017).

55. Leong, D. P. et al. Prognostic value of grip strength: findings from the Prospective Urban Rural Epidemiology (PURE) study. Lancet 386, 266–273 (2015).

56. Wang, Y.-C. et al. Synergistic effect of low handgrip strength and malnutrition on 4-year all-cause mortality in older males: A prospective longitudinal cohort study. Arch. Gerontol. Geriatr. 83, 217–222 (2019).

57. Eekhoff, E. M. W. et al. Relative importance of four functional measures as predictors of 15-year mortality in the older Dutch population. BMC Geriatr. 19, 92 (2019).

58. Whitney, D. G. & Peterson, M. D. The association between differing grip strength measures and mortality and cerebrovascular event in older adults: National Health and Aging Trends Study. Front. Physiol. 9, 1871 (2018).

59. Karlsen, T., Nauman, J., Dalen, H., Langhammer, A. & Wisløff, U. The combined association of skeletal muscle strength and physical activity on mortality in older women: The HUNT2 Study. Mayo Clin. Proc. 92, 710–718 (2017).

60. Bae, E.-J., Park, N.-J., Sohn, H.-S. & Kim, Y.-H. Handgrip strength and all-cause mortality in middle-aged and older Koreans. Int. J. Environ. Res. Public Health 16, (2019).

61. Kim, Y. et al. The combination of cardiorespiratory fitness and muscle strength, and mortality risk. Eur. J. Epidemiol. 33, 953–964 (2018).

62. Granic, A. et al. Initial level and rate of change in grip strength predict all-cause mortality in very old adults. Age Ageing 46, 970–976 (2017).

63. Oksuzyan, A. et al. Handgrip strength and its prognostic value for mortality in Moscow, Denmark, and England. PLoS One 12, e0182684 (2017).

64. Sasaki, H., Kasagi, F., Yamada, M. & Fujita, S. Grip strength predicts cause-specific mortality in middle-aged and elderly persons. Am. J. Med. 120, 337–342 (2007).

65. García-Hermoso, A. et al. Muscular strength as a predictor of all-cause mortality in an apparently healthy population: a systematic review and meta-analysis of data from approximately 2 million men and women. Arch. Phys. Med. Rehabil. 99, 2100–2113.e5 (2018).

66. Rijk, J. M., Roos, P. R., Deckx, L., van den Akker, M. & Buntinx, F. Prognostic value of handgrip strength in people aged 60 years and older: A systematic review and meta-analysis. Geriatr. Gerontol. Int. 16, 5–20 (2016).

67. Bohannon, R. W. Hand-grip dynamometry predicts future outcomes in aging adults. J. Geriatr. Phys. Ther. 31, 3–10 (2008).

68. Dodds, R. M., Kuh, D., Sayer, A. A. & Cooper, R. Can measures of physical performance in mid-life improve the clinical prediction of disability in early old age? Findings from a British birth cohort study. Exp. Gerontol. 110, 118–124 (2018).

69. Olguín, T., Bunout, D., de la Maza, M. P., Barrera, G. & Hirsch, S. Admission handgrip strength predicts functional decline in hospitalized patients. Clin Nutr ESPEN 17, 28–32 (2017).

70. García-Peña, C. et al. Handgrip strength predicts functional decline at discharge in hospitalized male elderly: a hospital cohort study. PLoS One 8, e69849 (2013).

71. Hashimoto, S. et al. Preoperative hand-grip strength can be a predictor of stair ascent and descent ability after total knee arthroplasty in female patients. J. Orthop. Sci. 25, 167–172 (2020).

72. Di Monaco, M. et al. Handgrip strength is an independent predictor of functional outcome in hip-fracture women: a prospective study with 6-month follow-up. Medicine 94, e542 (2015).

73. Di Monaco, M. et al. Handgrip strength but not appendicular lean mass is an independent predictor of functional outcome in hip-fracture women: a short-term prospective study. Arch. Phys. Med. Rehabil. 95, 1719–1724 (2014).

74. McGrath, R. P. et al. The association between handgrip strength and diabetes on activities of daily living disability in older Mexican Americans. J. Aging Health 30, 1305–1318 (2018).

75. Cöster, M. E. et al. Physical function tests predict incident falls: A prospective study of 2969 men in the Swedish Osteoporotic Fractures in Men study. Scand. J. Public Health 48, 436–441 (2020).

76. Dufour, A. B., Kiel, D. P., Williams, S. A., Weiss, R. J. & Samelson, E. J. Risk factors for incident fracture in older adults with type 2 diabetes: The Framingham Heart Study. Diabetes Care 44, 1547–1555 (2021).

77. Rikkonen, T. et al. Long-term effects of functional impairment on fracture risk and mortality in postmenopausal women. Osteoporos. Int. 29, 2111–2120 (2018).

78. Miller Giles, L. C., Crotty, M., Harrison, J. E. & Andrews, G. R. A clinically relevant criterion for grip strength: relationship with falling in a sample of older adults. Nutr. Diet. 60, 248–252 (2003).

79. Dixon, W. G. et al. Low grip strength is associated with bone mineral density and vertebral fracture in women. Rheumatology 44, 642–646 (2005).

80. Finigan, J. et al. Risk factors for vertebral and nonvertebral fracture over 10 years: a population-based study in women. J. Bone Miner. Res. 23, 75–85 (2008).

81. Rouzi, A. A., Al-Sibiani, S. A., Al-Senani, N. S., Radaddi, R. M. & Ardawi, M.-S. M. Independent predictors of all osteoporosis-related fractures among healthy Saudi postmenopausal women: the CEOR Study. Bone 50, 713–722 (2012).

82. Sirola, J. et al. Grip strength may facilitate fracture prediction in perimenopausal women with normal BMD: a 15-year population-based study. Calcif. Tissue Int. 83, 93–100 (2008).

83. Arges, K. et al. The Project Baseline Health Study: a step towards a broader mission to map human health. NPJ Digit. Med. 3, 84 (2020).

84. Califf, R. M. et al. Biological and clinical correlates of the patient health questionnaire-9: exploratory cross-sectional analyses of the baseline health study. BMJ Open 12, e054741 (2022).

85. McGrath, R. P., Kraemer, W. J., Snih, S. A. & Peterson, M. D. Handgrip strength and health in aging adults. Sports Med. 48, 1993–2000 (2018).

86. Germain, C. M., Vasquez, E., Batsis, J. A. & McQuoid, D. R. Sex, race and age differences in muscle strength and limitations in community dwelling older adults: Data from the Health and Retirement Survey (HRS). Arch. Gerontol. Geriatr. 65, 98–103 (2016).

87. Silva, A. M. et al. Ethnicity-related skeletal muscle differences across the lifespan. Am. J. Hum. Biol. 22, 76–82 (2010).

88. McFadden, D. & Bracht, M. S. Sex and race differences in the relative lengths of metacarpals and metatarsals in human skeletons. Early Hum. Dev. 85, 117–124 (2009).

89. VanderWeele, T. J. & Robinson, W. R. On the causal interpretation of race in regressions adjusting for confounding and mediating variables. Epidemiology 25, 473–484 (2014).

90. Wilson, D. et al. Frailty is associated with neutrophil dysfunction which is correctable with phosphoinositol-3-kinase inhibitors. J. Gerontol. A Biol. Sci. Med. Sci. 75, 2320–2325 (2020).

91. Beyer, I. et al. Inflammation-related muscle weakness and fatigue in geriatric patients. Exp. Gerontol. 47, 52–59 (2012).

92. Kim, S. W., Lee, H. A. & Cho, E.-H. Low handgrip strength is associated with low bone mineral density and fragility fractures in postmenopausal healthy Korean women. J. Korean Med. Sci. 27, 744–747 (2012).

93. Kritz-Silverstein, D. & Barrett-Connor, E. Grip strength and bone mineral density in older women. J. Bone Miner. Res. 9, 45–51 (1994).

94. Li, Y.-Z. et al. Low grip strength is a strong risk factor of osteoporosis in postmenopausal women. Orthop. Surg. 10, 17–22 (2018).

95. Logan, S. et al. Chronic joint pain and handgrip strength correlates with osteoporosis in mid-life women: a Singaporean cohort. Osteoporos. Int. 28, 2633–2643 (2017).

96. Miyakoshi, N. et al. Comparison of spinal alignment, muscular strength, and quality of life between women with postmenopausal osteoporosis and healthy volunteers. Osteoporos. Int. 28, 3153–3160 (2017).

97. Nagai, A. et al. Relations between quantitative ultrasound assessment of calcaneus and grip and key pinch power in Japanese mountain village residents. J. Orthop. Surg. 25, 2309499017690321 (2017).

98. Tachiki, T. et al. Muscle strength is associated with bone health independently of muscle mass in postmenopausal women: the Japanese population-based osteoporosis study. J. Bone Miner. Metab. 37, 53–59 (2019).

99. Kärkkäinen, M. et al. Physical tests for patient selection for bone mineral density measurements in postmenopausal women. Bone 44, 660–665 (2009).

100. Ma, Y. et al. Muscle strength rather than muscle mass is associated with osteoporosis in older Chinese adults. J. Formos. Med. Assoc. 117, 101–108 (2018).

101. McGrath, R. P., Kraemer, W. J., Vincent, B. M., Hall, O. T. & Peterson, M. D. Muscle strength is protective against osteoporosis in an ethnically diverse sample of adults. J. Strength Cond. Res. 31, 2586–2589 (2017).

102. Lee, K. Weight underestimation and weight nonregulation behavior may be related to weak grip strength. Nutr. Res. 87, 41–48 (2021).

103. Gordon, B. R., McDowell, C. P., Lyons, M. & Herring, M. P. Associations between grip strength and generalized anxiety disorder in older adults: Results from the Irish longitudinal study on ageing. J. Affect. Disord. 255, 136–141 (2019).

104. van Milligen, B. A., Lamers, F., de Hoop, G. T., Smit, J. H. & Penninx, B. W. J. H. Objective physical functioning in patients with depressive and/or anxiety disorders. J. Affect. Disord. 131, 193–199 (2011).

105. Hotham, E., Haberfield, M., Hillier, S., White, J. M. & Todd, G. Upper limb function in children with attention-deficit/hyperactivity disorder (ADHD). J. Neural Transm. 125, 713–726 (2018).

106. World Health Organization. Measuring Health and Disability: Manual for WHO Disability Assessment Schedule WHODAS 2.0. (World Health Organization, 2010).

107. Nelson, H. D., Nevitt, M. C., Scott, J. C., Stone, K. L. & Cummings, S. R. Smoking, alcohol, and neuromuscular and physical function of older women. Study of Osteoporotic Fractures Research Group. J.A.M.A. 272, 1825–1831 (1994).

108. Quan, S., Jeong, J.-Y. & Kim, D.-H. The relationship between smoking, socioeconomic status and grip strength among community-dwelling elderly men in Korea: Hallym Aging Study. Epidemiol. Health 35, e2013001 (2013).

109. Strand, B. H., Mishra, G., Kuh, D., Guralnik, J. M. & Patel, K. V. Smoking history and physical performance in midlife: results from the British 1946 birth cohort. J. Gerontol. A Biol. Sci. Med. Sci. 66, 142–149 (2011).

110. Tufan, A. et al. Low glomerular filtration rate as an associated risk factor for sarcopenic muscle strength: is creatinine or cystatin C-based estimation more relevant? Aging Male 20, 110–114 (2017).

111. Dalrymple, L. S. et al. Kidney function and prevalent and incident frailty. Clin. J. Am. Soc. Nephrol. 8, 2091–2099 (2013).

112. Corsonello, A. et al. Estimated glomerular filtration rate and functional status among older people: A systematic review. Eur. J. Intern. Med. 56, 39–48 (2018).

113. Mayeda, E. R. & Glymour, M. M. The obesity paradox in survival after cancer diagnosis: tools for evaluation of potential bias. Cancer Epidemiol. Biomarkers Prev. 26, 17–20 (2017).

114. Whitcomb, B. W., Schisterman, E. F., Perkins, N. J. & Platt, R. W. Quantification of collider-stratification bias and the birthweight paradox. Paediatr. Perinat. Epidemiol. 23, 394–402 (2009).

115. Munafò, M. R., Tilling, K., Taylor, A. E., Evans, D. M. & Davey Smith, G. Collider scope: when selection bias can substantially influence observed associations. Int. J. Epidemiol. 47, 226–235 (2018).

116. Holmberg, M. J. & Andersen, L. W. Collider bias. J.A.M.A. 327, 1282–1283 (2022).

117. Gubelmann, C., Vollenweider, P. & Marques-Vidal, P. Association of grip strength with cardiovascular risk markers. Eur. J. Prev. Cardiol. 24, 514–521 (2017).

118. Li, D. et al. Relative handgrip strength is inversely associated with metabolic profile and metabolic disease in the general population in China. Front. Physiol. 9, 59 (2018).

119. Lee, W.-J., Peng, L.-N., Chiou, S.-T. & Chen, L.-K. Relative handgrip strength is a simple indicator of cardiometabolic risk among middle-aged and older people: a nationwide population-based study in Taiwan. PLoS One 11, e0160876 (2016).

120. Hernan, M. A. & Robins, J. M. Causal Inference: What If. (Chapman & Hall/CRC, 2020).

121. United States Census Bureau. QuickFacts: United States.

122. Azur, M. J., Stuart, E. A., Frangakis, C. & Leaf, P. J. Multiple imputation by chained equations: what is it and how does it work? Int. J. Methods Psychiatr. Res. 20, 40–49 (2011).

123. Chen, Q. & Wang, S. Variable selection for multiply-imputed data with application to dioxin exposure study. Stat. Med. 32, 3646–3659 (2013).

124. Rubin, D. B. Multiple Imputation for Nonresponse in Surveys. (Wiley, 1987). doi:10.1002/9780470316696.

125. van Buuren, S. & Groothuis-Oudshoorn, K. mice: Multivariate Imputation by Chained Equations in R. J. Stat. Softw. 45, 1–67 (2011).

126. R Core Team. R: A language and environment for statistical computing. R Foundation for Statistical Computing, Vienna, Austria, 2020. Available at: https://www.R-project.org/.

127. Wickham, H. ggplot2: Elegant Graphics for Data Analysis. (Springer, 2016).

